# Internal and External Validation of Machine Learning Algorithms Versus FINDRISC for Incident Type 2 Diabetes: A Transparent, Explainable Benchmark Using SHAP

**DOI:** 10.1101/2025.09.05.25335151

**Authors:** Parsa Amirian, Mahsa Zarpoosh

**Author notes:** Corresponding author, Address: Kermanshah University of Medical Sciences (KUMS), Shahid Beheshti Boulevard, Kermanshah, Iran. Address: Kermanshah University of Medical Sciences(KUMS), Shahid Beheshti Boulevard, Kermanshah, Iran.

## Abstract

**Background:** Type 2 diabetes mellitus (T2DM) affects almost half a billion people, and the projected cost is $2.25 trillion by 2030; early detection strategies are needed for prevention. However, Finnish Diabetes Risk Score (FINDRISC) enables questionnaire-based risk assessment; its linearity and lack of interpretability limit predictive power and generalizability across diverse populations. Machine learning (ML) models are the next-generation prediction tools, but require rigorous benchmarking, external validation, and explainability to be clinically trusted.

**Methods:** In the current prospective cohort (n=9,171, 7.1 years follow-up), we compared six supervised ML models, three anomaly detectors, and a stacking ensemble against FINDRISC for T2DM incidence, using harmonized, calibrated pipelines and internal and external validation in US (NHANES) and PIMA Indian populations. External validations included reduced (7- and 3-variable) models, and explainability was assessed with SHAP.

**Results:** ML models, particularly neural networks and stacking, achieved superior internal discrimination (ROC AUC up to 0.87 vs. FINDRISC 0.70), with stacking ensemble recall of 0.81. In reduced-variable external validations, ML models maintained robust performance (AUCs > 0.76), and strikingly, the isolation forest anomaly detector excelled in US data. Sensitivity analysis demonstrated that without laboratory data, FINDRISC still matches or exceeds ML, thereby preserving its practical role in non-laboratory settings. SHAP analysis consistently identified FBS, BMI, and age as main predictors, promoting interpretability.

**Conclusions:** Harmonized ML models, when externally validated, substantially improve traditional risk scores for T2DM prediction, particularly when laboratory data are available. Transparent analytics and an open-source online calculator support global clinical deployment. This work substantially advances precision prevention in T2DM through explainable, portable prediction.

## Introduction

Type 2 diabetes (T2DM) is one of the leading global health challenges; it affects almost half a billion people worldwide. By 2050, its prevalence is projected to rise by 45% (1). T2DM is associated with morbidities, namely cardiovascular diseases (CVDs), kidney failures, neuropathies, and increased mortalities among affected populations. It also represents a significant financial burden for countries, with an estimated total cost of T2DM reaching $2.25 trillion by 2030 (1, 2). Early prediction of T2DM is invaluable because early response and treatment could mitigate or diminish its complications, including myocardial infarction (MI), hyperglycemic crises, stroke, amputations, and renal diseases substantially. Moreover, as a global health problem, the incidence of T2DM is increasing; therefore, as advocated by leading diabetes and digital health recommendations for creating readily deployable risk prediction models, our work supports precision prevention across diverse global populations. (1,3).

The Finnish Diabetes Risk Score (FINDRISC) is one of the established non-laboratory systems used to predict T2DM. It is a questionnaire developed to identify individuals at risk of developing T2DM; the main components include age, body mass index (BMI), physical activity, and waist circumference (4). Improved risk prediction tools that account for complex patterns in data and can be reliably deployed across diverse populations are urgently needed; however, FINDRISC does not capture nonlinear relationships or interactions between predictors. Furthermore, there are significant gaps in its interpretability.

Mainstream supervised machine learning (ML) classifiers, including random forest (RF), logistic regression (LR), and support vector machine (SVM), have been utilized to predict the incidence of T2DM (5-7). While ML approaches such as RF, LR, and SVM have demonstrated potential for T2DM risk prediction, prior studies often lack external validation, feature-limited candidate predictors, and rarely assess calibration or clinical utility; in addition, reproducible analytic pipelines and estimates of uncertainty are seldom provided (5). Notably, 85% of studies only rely on the mentioned classifiers to predict the incidence of T2DM (8). Generally speaking, the incidence of new diabetes cases in population-based cohorts is often low and can be considered anomalous events (9). Based on this fact, we have added three anomaly detection models, local outlier factor (LOF), one-class SVM (OC-SVM), and isolation forest (IF) to our main ML models. In addition to deriving reliable insights from our T2DM dataset, we trained a stacking ensemble model to leverage the strengths of the base models. We also validated our models not only within the internal cohort but also externally in two ethnically distinct populations that are publicly available to researchers: a representative US cohort, the National Health and Nutrition Examination Survey (NHANES), and the Indian PIMA women’s cohort.

This study aimed to perform head-to-head benchmarking of standard classifiers, anomaly detection frameworks, and an ensemble against a well-established clinical scoring system (FINDRISC) as a clinical reference, and to validate models in ethnically diverse external cohorts. Additionally, we offer rigorously harmonized pipelines for T2DM risk prediction, complete with an online risk calculator. To enhance the robustness and credibility of any observed improvements, we conducted sensitivity analyses comparing models. Additionally, to make our models more understandable and transparent, we calculated Shapley additive explanations (SHAP) values. In this study, we benchmarked ML models against FINDRISC internally and externally validated them. Such comprehensive analytic transparency and clinical utility, in identifying a robust, transparent, and globally applicable T2DM risk prediction tool, move beyond conventional approaches.

## Methods

### Study Design, Population, and Data Sources

We utilized data from our internal cohort, the Ravansar Non-Communicable Disease (RaNCD) prospective cohort, which comprises 10,000 recruited participants aged 35 to 70. Moreover, these participants reside in both urban and rural areas of Ravansar County in the northwestern part of Kermanshah Province (10). The first phase of the cohort commenced in 2015, and as of 2023, six follow-up sessions have been completed. The data were obtained through various means, including anthropometry, physical examination, questionnaires, and laboratory tests. To maximize the available subjects, we included all the original 10047 participants. We used different imputation methods based on data exploration (nature of the data to be either categorical or continuous). For categorical variables, we employed the mode imputation method, which resulted in 34 instances being imputed. Two non-binary categorical variables (smoking status and T2DM prevalence) had 164 instances of missing data; in these cases, we utilized the proportional imputation method. Finally, in the case of continuous variables, 308 data points were missing; therefore, we used the mean imputation method for those variables. We did not detect any patterns in our missing data, and they most likely were completely random. To boost the performance of our ML models, we employed a clipping approach for variables with outliers. We converted all continuous variables into z-scores. Subsequently, any data point that was more than three standard deviations (SD) from the mean (z-score > 3 or < -3) was considered an outlier. Across six variables, 712 instances were clipped. Out of our initial 10047 cohort participants, 876 (8.72%) were already diagnosed with T2DM; as a result, we excluded those participants and started with 9171 entrants. For exploratory external validation of our trained ML models, we used two publicly available T2DM datasets: 1. National Health and Nutrition Examination Survey (NHANES) 2013-2014 (Kaggle: https://www.kaggle.com/datasets/cdc/national-health-and-nutrition-examination-survey) and 2. PIMA Indian women T2DM dataset (Kaggle: https://www.kaggle.com/datasets/uciml/pima-indians-diabetes-database) (11,12). Across all cohorts, we removed every observation with an FBS more than 126. Due to this rule, no one was removed from our internal cohort (after excluding prevalence cases); nonetheless, 4270 cases were excluded from the original 10176 NHANES participants. To ensure maximum generalizability, we adopted a different approach in addressing the missing data in the external cohorts. We avoided any synthesized data and removed all participants with missing data in the selected variables. Ultimately, we removed 57 observations due to missing BMI, 499 observations due to missing socioeconomic status quartile (SESq), 1085 observations due to missing CVD, and 122 observations due to missing T2DM status. Finally, 80 observations were removed because of missing T2DM familial history. As a result, 6,113 participants were removed from the analysis, and 4,062 NHANES participants were included in the external validation. The PIMA Indian women database had no missing values; however, 288 participants had an FBS greater than 126 and were therefore removed from the analysis. The formal analysis was conducted on 480 participants (see Table 1 and Figure 1 for a schematic of the whole analytic workflow). All continuous variables were converted to z-scores using the internal cohort training set means/SD exclusively, to prevent information leakage into test/external sets; in addition, no clipping was performed across external cohorts, and they were stratified randomly into 80% train and 20% test sets.

**Table 1.**
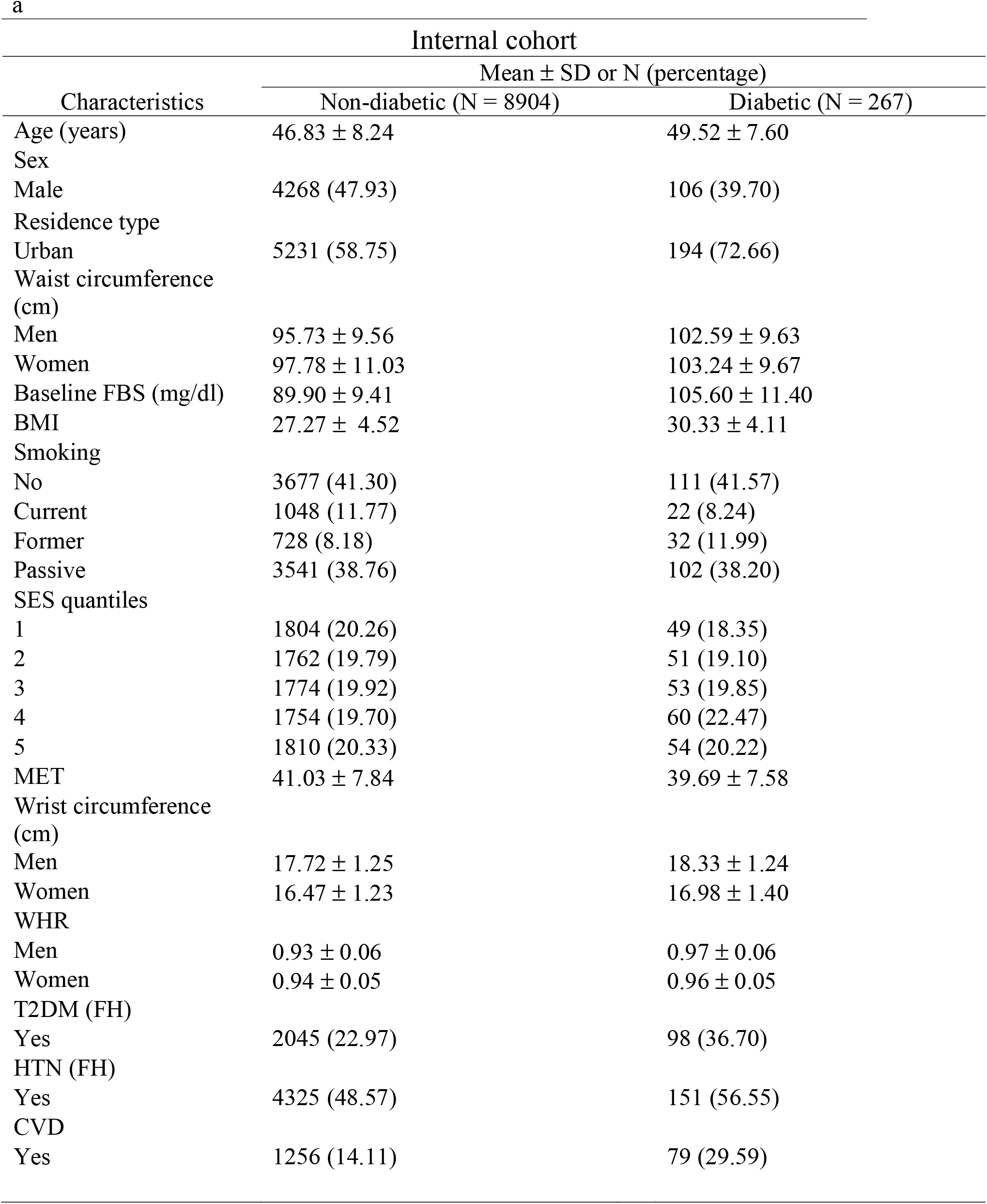
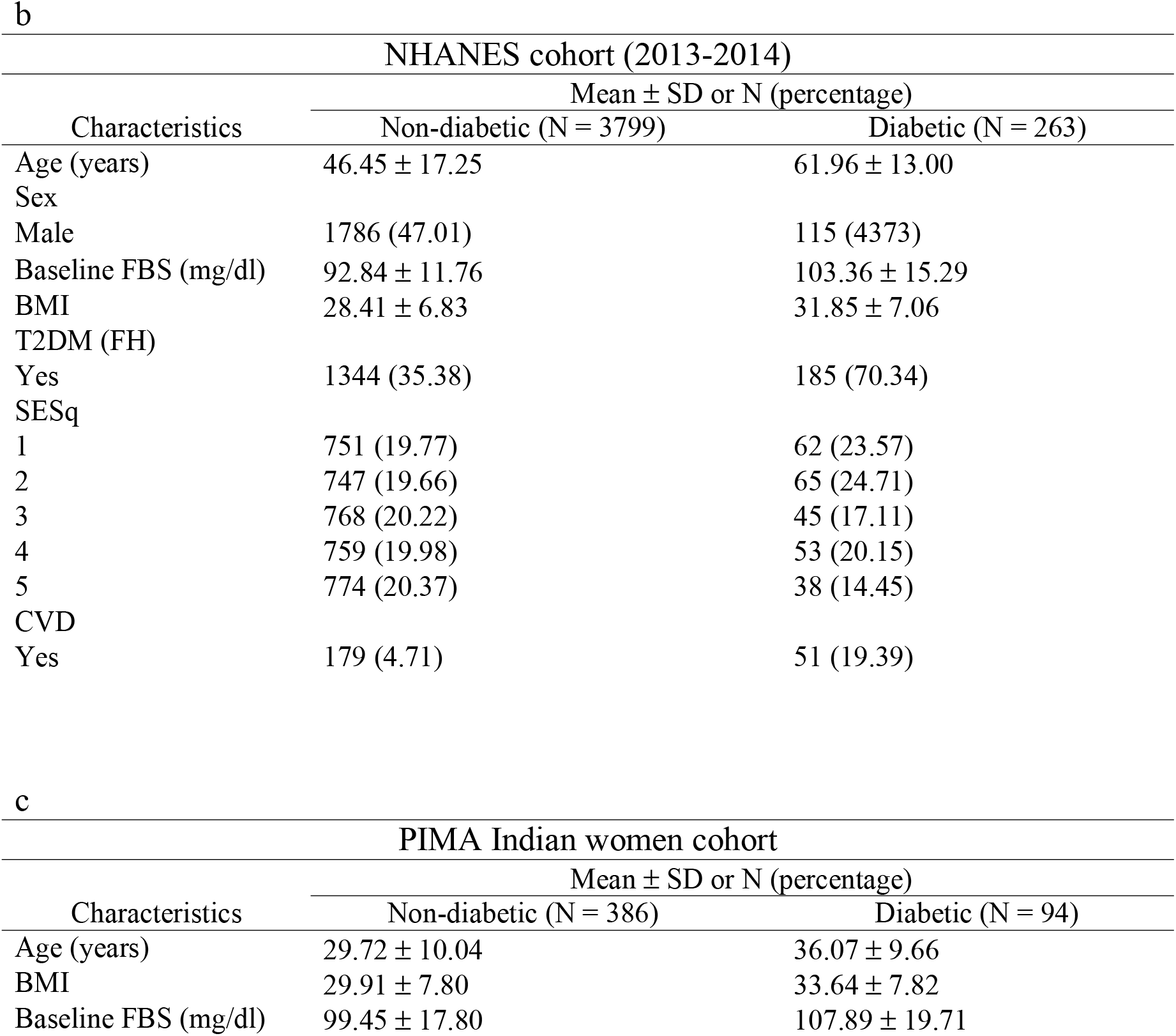
Demographic characteristics of the participants in the cohorts. (a) Internal cohort (b), NHANES cohort (c), PIMA Indian women cohort.

Internal cohort
| Characteristics | Mean ± SD or N (percentage) | |
| --- | --- | --- |
|  | Non-diabetic (N = 8904) | Diabetic (N = 267) |
| Age (years) | 46.83 ± 8.24 | 49.52 ± 7.60 |
| Sex |  |  |
| Male | 4268 (47.93) | 106 (39.70) |
| Residence type |  |  |
| Urban | 5231 (58.75) | 194 (72.66) |
| Waist circumference (cm) |  |  |
| Men | 95.73 ± 9.56 | 102.59 ± 9.63 |
| Women | 97.78 ± 11.03 | 103.24 ± 9.67 |
| Baseline FBS (mg/dl) | 89.90 ± 9.41 | 105.60 ± 11.40 |
| BMI | 27.27 ± 4.52 | 30.33 ± 4.11 |
| Smoking |  |  |
| No | 3677 (41.30) | 111 (41.57) |
| Current | 1048 (11.77) | 22 (8.24) |
| Former | 728 (8.18) | 32 (11.99) |
| Passive | 3541 (38.76) | 102 (38.20) |
| SES quantiles |  |  |
| 1 | 1804 (20.26) | 49 (18.35) |
| 2 | 1762 (19.79) | 51 (19.10) |
| 3 | 1774 (19.92) | 53 (19.85) |
| 4 | 1754 (19.70) | 60 (22.47) |
| 5 | 1810 (20.33) | 54 (20.22) |
| MET | 41.03 ± 7.84 | 39.69 ± 7.58 |
| Wrist circumference (cm) |  |  |
| Men | 17.72 ± 1.25 | 18.33 ± 1.24 |
| Women | 16.47 ± 1.23 | 16.98 ± 1.40 |
| WHR |  |  |
| Men | 0.93 ± 0.06 | 0.97 ± 0.06 |
| Women | 0.94 ± 0.05 | 0.96 ± 0.05 |
| T2DM (FH) |  |  |
| Yes | 2045 (22.97) | 98 (36.70) |
| HTN (FH) |  |  |
| Yes | 4325 (48.57) | 151 (56.55) |
| CVD |  |  |
| Yes | 1256 (14.11) | 79 (29.59) |

| NHANES cohort (2013-2014) |  |  |
| --- | --- | --- |
| Characteristics | Mean ± SD or N (percentage) | |
|  | Non-diabetic (N = 3799) | Diabetic (N = 263) |
| Age (years) | 46.45 ± 17.25 | 61.96 ± 13.00 |
| Sex |  |  |
| Male | 1786 (47.01) | 115 (43.73) |
| Baseline FBS (mg/dl) | 92.84 ± 11.76 | 103.36 ± 15.29 |
| BMI | 28.41 ± 6.83 | 31.85 ± 7.06 |
| T2DM (FH) |  |  |
| Yes | 1344 (35.38) | 185 (70.34) |
| SESq |  |  |
| 1 | 751 (19.77) | 62 (23.57) |
| 2 | 747 (19.66) | 65 (24.71) |
| 3 | 768 (20.22) | 45 (17.11) |
| 4 | 759 (19.98) | 53 (20.15) |
| 5 | 774 (20.37) | 38 (14.45) |
| CVD |  |  |
| Yes | 179 (4.71) | 51 (19.39) |

| PIMA Indian women cohort |  |  |
| --- | --- | --- |
| Characteristics | Mean ± SD or N (percentage) | |
|  | Non-diabetic (N = 386) | Diabetic (N = 94) |
| Age (years) | 29.72 ± 10.04 | 36.07 ± 9.66 |
| BMI | 29.91 ± 7.80 | 33.64 ± 7.82 |
| Baseline FBS (mg/dl) | 99.45 ± 17.80 | 107.89 ± 19.71 |

**Figure 1.**
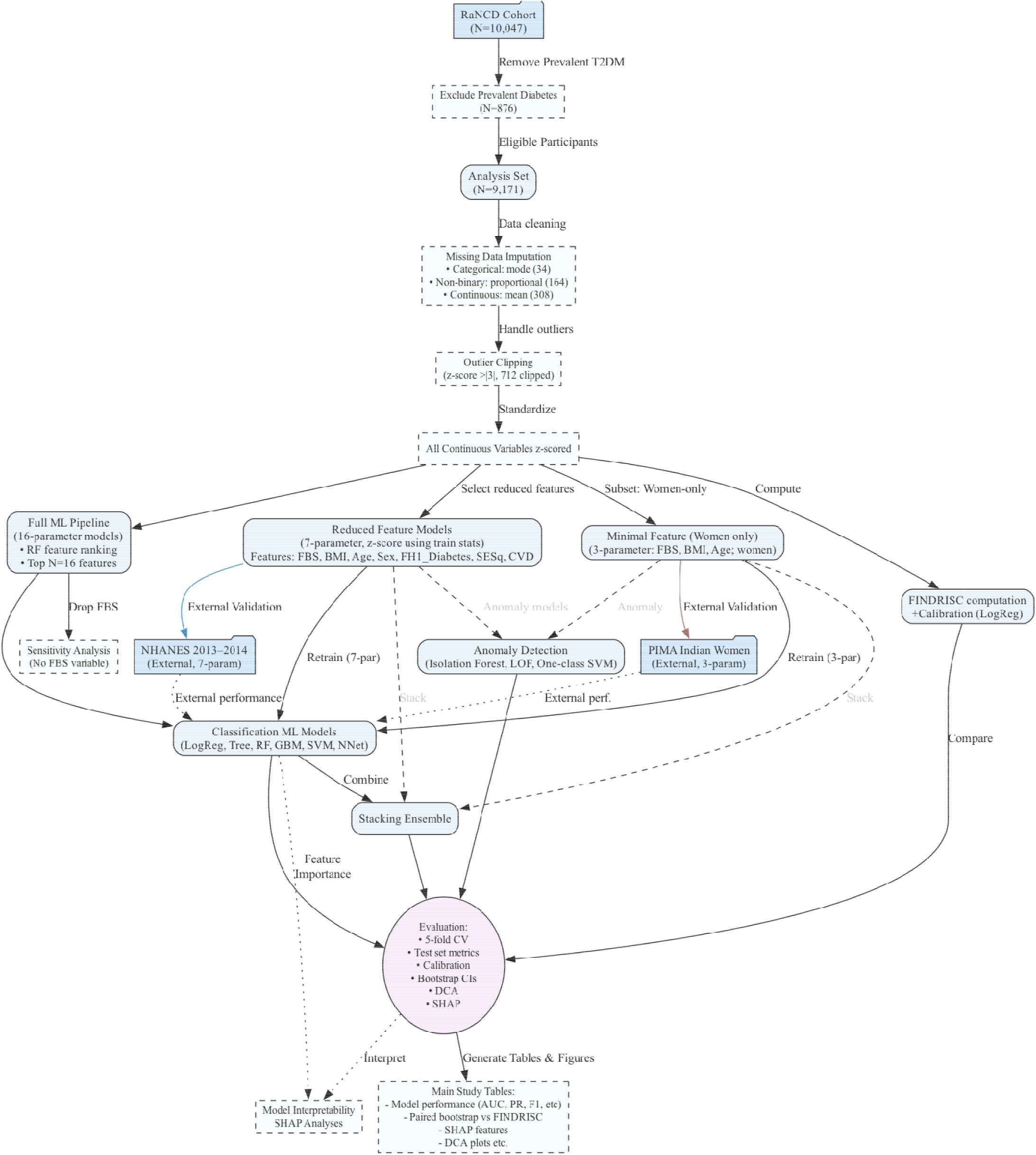
Study Design and Pipeline Overview.

### FINDRISC score

By employing FINDRISC’s original algorithm, we calculated the total score of each entrant by assigning integer points for each domain and then summing them up (13). The dataset did not contain self-reported exercise time for each day; therefore, we used our MET variable, which is the sum of all activity MET-hours over 24 hours. By using this variable and performing some calculations, we set 30 as our FINDRISC cutoff for exercise. Additionally, instead of a history of elevated blood glucose, which FINDRISC inquires about, we utilized the baseline fasting blood sugar (FBS) variable. We scored 5 points for participants with FBS higher than 126 milligram/ deciliter (mg/dl), but due to prior exclusion criteria of FBS more than 126, nobody achieved 5 points in this domain. Lastly, the part on dietary habits of participants was missing from our dataset; therefore, we omitted this component entirely and assigned all members zero points. Ultimately, our maximum possible FINDRISC score was lower than that in the original algorithm. After calculating the raw score of each member, we fitted a univariable logistic regression model using the raw score as the sole predictor on 80% of the dataset. We applied it to the remaining 20%. We applied the original FINDRISC cutoff of 15 points or more to classify participants as high or low risk. Table 1 of the S1 file displays the distribution of FINDRISC scores across the internal cohort.

### Features

We started with 27 potential predictor variables, and using a random forest classifier applied to the complete training set, we ranked the features from most to least important for the main internal analysis (Figure 1 S1 file) and sensitivity analysis (Figure 10 S1 file). The sorted features in main analysis are as follows: FBS, BMI, waist circumference, MET, age, waist-to-hip ratio (WHR), wrist circumference, SESq, familial history of T2DM, type of residence (urban or rural), history of cardiovascular diseases (CVD), smoking status (current, former, passive, none-smoker), familial history of hypertension (HTN), sex, anti-hypertensive drug history, familial history of cardiac diseases, familial history of stroke, HTN history, familial history of myocardial infarction (MI), history of gastroesophageal reflux disease (GERD), history of blood in stool, history of statin prescription, diagnosed with osteoporosis (yes or no), history of rheumatoid arthritis (RA), history of weight loss, alcohol consumption (yes or no), muscle weakness, and history of anti-triglyceride drug prescriptions. After carefully evaluating the NHANES dataset, we were able to map the definitions of 7 variables to align with those used in the internal Ravansar cohort. Table 2 of the S1 file summarizes the mapping of key features and the outcome variable. In our internal dataset, socioeconomic status was measured using a quintile ranking variable (SESq), with values from 1 (lowest SES) to 5 (highest SES). In NHANES, SES was represented by the family income-to-poverty ratio (INDFMPIR), a continuous variable. We mapped INDFMPIR into quintiles corresponding to the SESq scale, such that the lowest quintile of INDFMPIR was assigned a value of 1, and the highest quintile a value of 5.

**Table 2.**
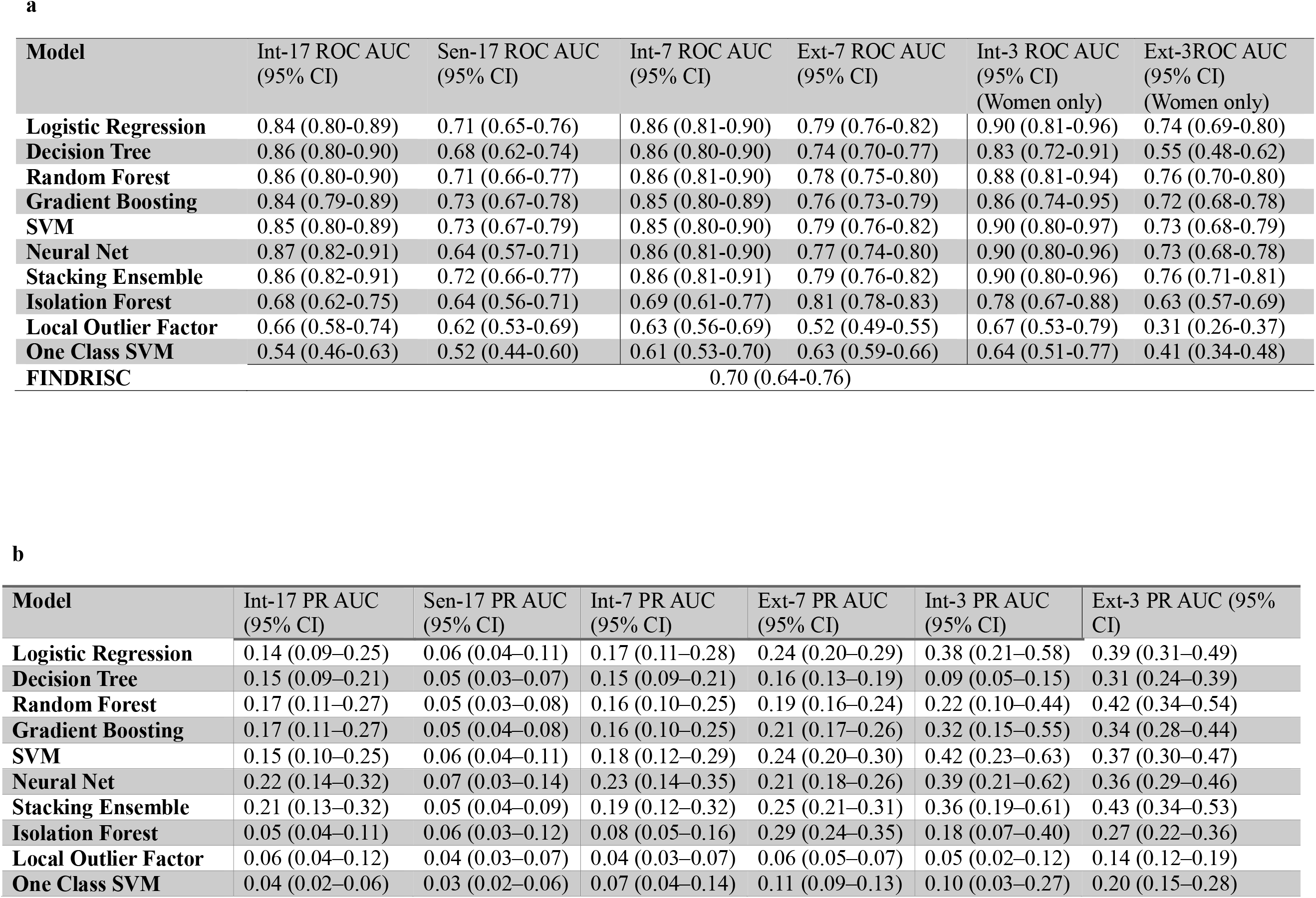
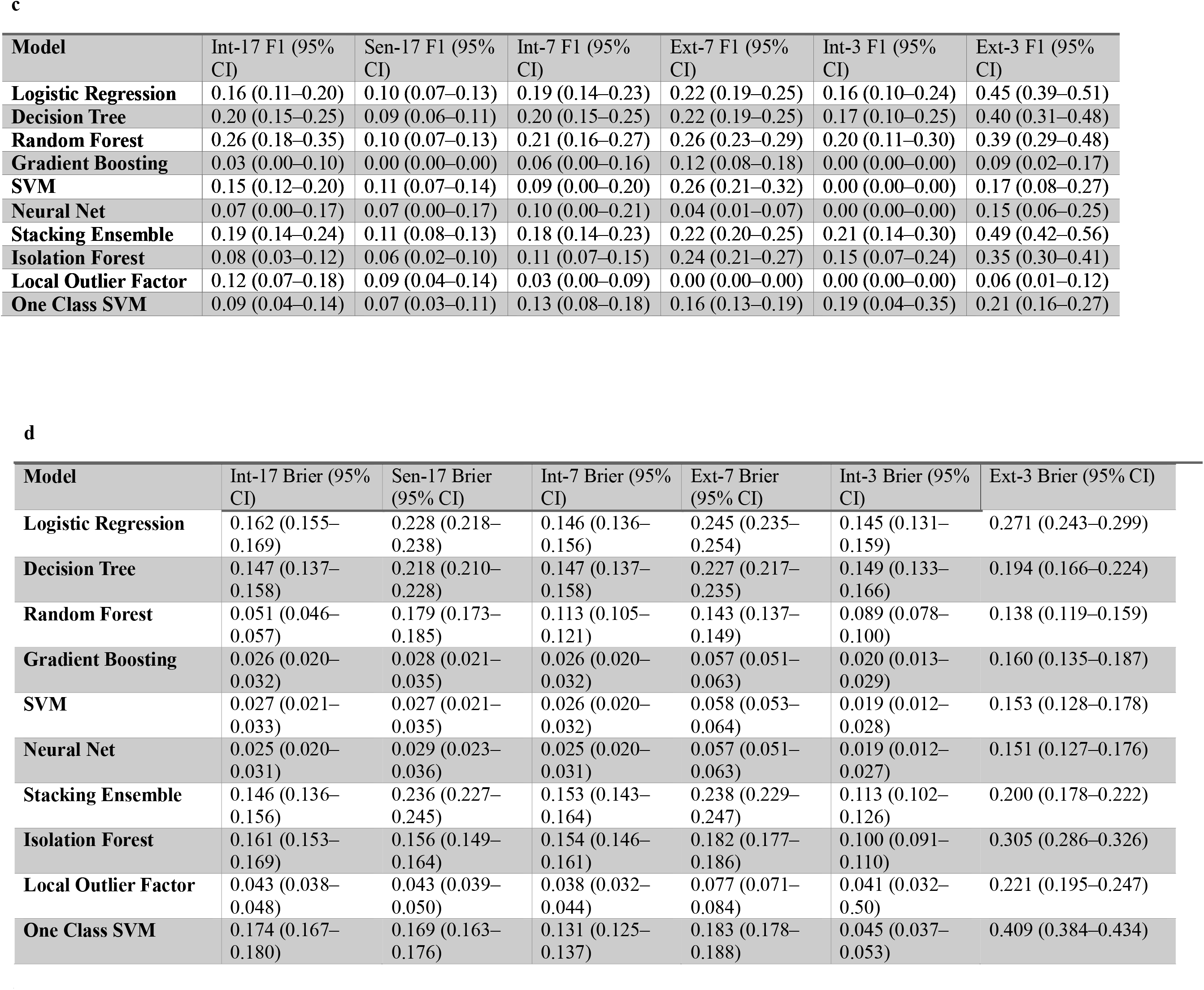
Comparison of all (a) ROC AUCs, (b) PR AUCs, (c) F1, and (d) Brier scores of different models and their 95% confidence interval (CI) across all cohorts.

| Model | Int-17 ROC AUC<br>(95% CI) | Sen-17 ROC AUC<br>(95% CI) | Int-7 ROC AUC<br>(95% CI) | Ext-7 ROC AUC<br>(95% CI) | Int-3 ROC AUC<br>(95% CI)<br>(Women only) | Ext-3 ROC AUC<br>(95% CI)<br>(Women only) |
| --- | --- | --- | --- | --- | --- | --- |
| **Logistic Regression** | 0.84 (0.80-0.89) | 0.71 (0.65-0.76) | 0.86 (0.81-0.90) | 0.79 (0.76-0.82) | 0.90 (0.81-0.96) | 0.74 (0.69-0.80) |
| **Decision Tree** | 0.86 (0.80-0.90) | 0.68 (0.62-0.74) | 0.86 (0.80-0.90) | 0.74 (0.70-0.77) | 0.83 (0.72-0.91) | 0.55 (0.48-0.62) |
| **Random Forest** | 0.86 (0.80-0.90) | 0.71 (0.66-0.77) | 0.86 (0.81-0.90) | 0.78 (0.75-0.80) | 0.88 (0.81-0.94) | 0.76 (0.70-0.80) |
| **Gradient Boosting** | 0.84 (0.79-0.89) | 0.73 (0.67-0.78) | 0.85 (0.80-0.89) | 0.76 (0.73-0.79) | 0.86 (0.74-0.95) | 0.72 (0.68-0.78) |
| **SVM** | 0.85 (0.80-0.89) | 0.73 (0.67-0.79) | 0.85 (0.80-0.90) | 0.79 (0.76-0.82) | 0.90 (0.80-0.97) | 0.73 (0.68-0.79) |
| **Neural Net** | 0.87 (0.82-0.91) | 0.64 (0.57-0.71) | 0.86 (0.81-0.90) | 0.77 (0.74-0.80) | 0.90 (0.80-0.96) | 0.73 (0.68-0.78) |
| **Stacking Ensemble** | 0.86 (0.82-0.91) | 0.72 (0.66-0.77) | 0.86 (0.81-0.91) | 0.79 (0.76-0.82) | 0.90 (0.80-0.96) | 0.76 (0.71-0.81) |
| **Isolation Forest** | 0.68 (0.62-0.75) | 0.64 (0.56-0.71) | 0.69 (0.61-0.77) | 0.81 (0.78-0.83) | 0.78 (0.67-0.88) | 0.63 (0.57-0.69) |
| **Local Outlier Factor** | 0.66 (0.58-0.74) | 0.62 (0.53-0.69) | 0.63 (0.56-0.69) | 0.52 (0.49-0.55) | 0.67 (0.53-0.79) | 0.31 (0.26-0.37) |
| **One Class SVM** | 0.54 (0.46-0.63) | 0.52 (0.44-0.60) | 0.61 (0.53-0.70) | 0.63 (0.59-0.66) | 0.64 (0.51-0.77) | 0.41 (0.34-0.48) |
| **FINDRISC** | 0.70 (0.64-0.76) |  |  |  |  |  |

b
| Model | Int-17 PR AUC<br>(95% CI) | Sen-17 PR AUC<br>(95% CI) | Int-7 PR AUC<br>(95% CI) | Ext-7 PR AUC<br>(95% CI) | Int-3 PR AUC<br>(95% CI) | Ext-3 PR AUC (95%<br>CI) |
| --- | --- | --- | --- | --- | --- | --- |
| **Logistic Regression** | 0.14 (0.09–0.25) | 0.06 (0.04–0.11) | 0.17 (0.11–0.28) | 0.24 (0.20–0.29) | 0.38 (0.21–0.58) | 0.39 (0.31–0.49) |
| **Decision Tree** | 0.15 (0.09–0.21) | 0.05 (0.03–0.07) | 0.15 (0.09–0.21) | 0.16 (0.13–0.19) | 0.09 (0.05–0.15) | 0.31 (0.24–0.39) |
| **Random Forest** | 0.17 (0.11–0.27) | 0.05 (0.03–0.08) | 0.16 (0.10–0.25) | 0.19 (0.16–0.24) | 0.22 (0.10–0.44) | 0.42 (0.34–0.54) |
| **Gradient Boosting** | 0.17 (0.11–0.27) | 0.05 (0.04–0.08) | 0.16 (0.10–0.25) | 0.21 (0.17–0.26) | 0.32 (0.15–0.55) | 0.34 (0.28–0.44) |
| **SVM** | 0.15 (0.10–0.25) | 0.06 (0.04–0.11) | 0.18 (0.12–0.29) | 0.24 (0.20–0.30) | 0.42 (0.23–0.63) | 0.37 (0.30–0.47) |
| **Neural Net** | 0.22 (0.14–0.32) | 0.07 (0.03–0.14) | 0.23 (0.14–0.35) | 0.21 (0.18–0.26) | 0.39 (0.21–0.62) | 0.36 (0.29–0.46) |
| **Stacking Ensemble** | 0.21 (0.13–0.32) | 0.05 (0.04–0.09) | 0.19 (0.12–0.32) | 0.25 (0.21–0.31) | 0.36 (0.19–0.61) | 0.43 (0.34–0.53) |
| **Isolation Forest** | 0.05 (0.04–0.11) | 0.06 (0.03–0.12) | 0.08 (0.05–0.16) | 0.29 (0.24–0.35) | 0.18 (0.07–0.40) | 0.27 (0.22–0.36) |
| **Local Outlier Factor** | 0.06 (0.04–0.12) | 0.04 (0.03–0.07) | 0.04 (0.03–0.07) | 0.06 (0.05–0.07) | 0.05 (0.02–0.12) | 0.14 (0.12–0.19) |
| **One Class SVM** | 0.04 (0.02–0.06) | 0.03 (0.02–0.06) | 0.07 (0.04–0.14) | 0.11 (0.09–0.13) | 0.10 (0.03–0.27) | 0.20 (0.15–0.28) |

c
| Model | Int-17 F1 (95% CI) | Sen-17 F1 (95% CI) | Int-7 F1 (95% CI) | Ext-7 F1 (95% CI) | Int-3 F1 (95% CI) | Ext-3 F1 (95% CI) |
| --- | --- | --- | --- | --- | --- | --- |
| **Logistic Regression** | 0.16 (0.11–0.20) | 0.10 (0.07–0.13) | 0.19 (0.14–0.23) | 0.22 (0.19–0.25) | 0.16 (0.10–0.24) | 0.45 (0.39–0.51) |
| **Decision Tree** | 0.20 (0.15–0.25) | 0.09 (0.06–0.11) | 0.20 (0.15–0.25) | 0.22 (0.19–0.25) | 0.17 (0.10–0.25) | 0.40 (0.31–0.48) |
| **Random Forest** | 0.26 (0.18–0.35) | 0.10 (0.07–0.13) | 0.21 (0.16–0.27) | 0.26 (0.23–0.29) | 0.20 (0.11–0.30) | 0.39 (0.29–0.48) |
| **Gradient Boosting** | 0.03 (0.00–0.10) | 0.00 (0.00–0.00) | 0.06 (0.00–0.16) | 0.12 (0.08–0.18) | 0.00 (0.00–0.00) | 0.09 (0.02–0.17) |
| **SVM** | 0.15 (0.12–0.20) | 0.11 (0.07–0.14) | 0.09 (0.00–0.20) | 0.26 (0.21–0.32) | 0.00 (0.00–0.00) | 0.17 (0.08–0.27) |
| **Neural Net** | 0.07 (0.00–0.17) | 0.07 (0.00–0.17) | 0.10 (0.00–0.21) | 0.04 (0.01–0.07) | 0.00 (0.00–0.00) | 0.15 (0.06–0.25) |
| **Stacking Ensemble** | 0.19 (0.14–0.24) | 0.11 (0.08–0.13) | 0.18 (0.14–0.23) | 0.22 (0.20–0.25) | 0.21 (0.14–0.30) | 0.49 (0.42–0.56) |
| **Isolation Forest** | 0.08 (0.03–0.12) | 0.06 (0.02–0.10) | 0.11 (0.07–0.15) | 0.24 (0.21–0.27) | 0.15 (0.07–0.24) | 0.35 (0.30–0.41) |
| **Local Outlier Factor** | 0.12 (0.07–0.18) | 0.09 (0.04–0.14) | 0.03 (0.00–0.09) | 0.00 (0.00–0.00) | 0.00 (0.00–0.00) | 0.06 (0.01–0.12) |
| **One Class SVM** | 0.09 (0.04–0.14) | 0.07 (0.03–0.11) | 0.13 (0.08–0.18) | 0.16 (0.13–0.19) | 0.19 (0.04–0.35) | 0.21 (0.16–0.27) |

| Model | Int-17 Brier (95% CI) | Sen-17 Brier (95% CI) | Int-7 Brier (95% CI) | Ext-7 Brier (95% CI) | Int-3 Brier (95% CI) | Ext-3 Brier (95% CI) |
| --- | --- | --- | --- | --- | --- | --- |
| **Logistic Regression** | 0.162 (0.155–0.169) | 0.228 (0.218–0.238) | 0.146 (0.136–0.156) | 0.245 (0.235–0.254) | 0.145 (0.131–0.159) | 0.271 (0.243–0.299) |
| **Decision Tree** | 0.147 (0.137–0.158) | 0.218 (0.210–0.228) | 0.147 (0.137–0.158) | 0.227 (0.217–0.235) | 0.149 (0.133–0.166) | 0.194 (0.166–0.224) |
| **Random Forest** | 0.051 (0.046–0.057) | 0.179 (0.173–0.185) | 0.113 (0.105–0.121) | 0.143 (0.137–0.149) | 0.089 (0.078–0.100) | 0.138 (0.119–0.159) |
| **Gradient Boosting** | 0.026 (0.020–0.032) | 0.028 (0.021–0.035) | 0.026 (0.020–0.032) | 0.057 (0.051–0.063) | 0.020 (0.013–0.029) | 0.160 (0.135–0.187) |
| **SVM** | 0.027 (0.021–0.033) | 0.027 (0.021–0.035) | 0.026 (0.020–0.032) | 0.058 (0.053–0.064) | 0.019 (0.012–0.028) | 0.153 (0.128–0.178) |
| **Neural Net** | 0.025 (0.020–0.031) | 0.029 (0.023–0.036) | 0.025 (0.020–0.031) | 0.057 (0.051–0.063) | 0.019 (0.012–0.027) | 0.151 (0.127–0.176) |
| **Stacking Ensemble** | 0.146 (0.136–0.156) | 0.236 (0.227–0.245) | 0.153 (0.143–0.164) | 0.238 (0.229–0.247) | 0.113 (0.102–0.126) | 0.200 (0.178–0.222) |
| **Isolation Forest** | 0.161 (0.153–0.169) | 0.156 (0.149–0.164) | 0.154 (0.146–0.161) | 0.182 (0.177–0.186) | 0.100 (0.091–0.110) | 0.305 (0.286–0.326) |
| **Local Outlier Factor** | 0.043 (0.038–0.048) | 0.043 (0.039–0.050) | 0.038 (0.032–0.044) | 0.077 (0.071–0.084) | 0.041 (0.032–0.50) | 0.221 (0.195–0.247) |
| **One Class SVM** | 0.174 (0.167–0.180) | 0.169 (0.163–0.176) | 0.131 (0.125–0.137) | 0.183 (0.178–0.188) | 0.045 (0.037–0.053) | 0.409 (0.384–0.434) |

Furthermore, no mapping was required for the PIMA Indian women dataset, as it was relatively small and the variables and outcome aligned with those in our internal cohort. Moreover, we used three predictors from the PIMA dataset, including age, BMI, and baseline FBS, as well as the outcome. To determine the optimal number of top features to retain (N) in the internal cohort, we evaluated N = 8, 12, 16, 20, and 24; for each N, we trained all seven baseline classifiers using stratified 5-fold cross-validation. Models with fewer than 16 features lost their discriminative power, while models with more than 16 predictors gained no additional power and were more prone to overfitting; as a result, we chose the top 16 features selected by RF feature importance (Table 2 S2 file).

### Data partitioning

Firstly, we dropped the label (T2DM incidence) to form the outcome column. Then, we split our dataset into an 80% training set and a 20% test set, and, to allow for reproducibility of the results, fixed the random state to 42. Similar to any other cohort with real-world data, our new T2DM incidence rates were extremely low (267 cases); this made our T2DM-positive class underrepresented. To address the mentioned class imbalance, we performed several exploratory runs using different methods, including the Synthetic Minority Over-sampling Technique (SMOTE), SMOTE combined with edited nearest neighbors (SMOTEENN), random oversampling, random undersampling, and class-weighting. We observed that the pure class-weighting method yielded the optimal results; therefore, we addressed class imbalance using the class-weighting method without data-level resampling. The intuitive reason might be due to the preservation of all real data; we kept important borderline examples, and because no synthetic artifacts were created, the models were able to generalize better under real-world data and the external cohorts. Class-weighting provided a robust and reproducible method to bias the models toward identifying new T2DM incidences without introducing fictional data to the models by simply penalizing mistakes on the minority class more heavily.

### Supervised classifiers

We trained six classifiers, including Decision Tree (DT), Gradient Boosting Machine (GBM), Neural Network (NN), SVM, LR, and RF, to predict new incidences of T2DM. Here is a brief breakdown of each model: DT models can be considered as large trees starting from their roots. They are non-parametric models that repeatedly separate the data at each node that maximizes a purity criterion. DT models are easy to understand, as if-then rules govern them; yet, they are prone to overfitting (14). GBM models are an ensemble of weak learners. When the model reiterates, a new tree is created and fitted to the negative gradient of a chosen loss function, therefore correcting residual errors (15).

On the other hand, NNs are feed-forward networks consisting of an input layer, hidden layers, and an output layer. These models are particularly advantageous in finding extremely complex relationships (16). The SVM’s primary function is to create a hyperplane that divides the feature space, separating two classes. SVM models are sturdy against high-dimensional data, but are computationally costly (17). LR models can be considered one of the simplest classifiers in the context of ML models; they predict labels by estimating the linear combination of predictors, and the resulting odds ratios indicate the direction and strength of their relationship (18). RF models are an ensemble of bootstrap-aggregated DTs; in the forest, each tree is grown on a random sample and a random subset of predictors within the data. They are highly resilient to overfitting, making them a good choice in the medical field (19). We performed a randomized grid search over pre□specified parameter distributions for each algorithm. Hyperparameter tuning for each model was performed using stratified five-fold cross-validation, with the mean ROC AUC across folds as the primary optimization metric. Random search was used to explore the parameter ranges specified in Note 1 of the S2 file. The best parameter sets were selected based on the highest mean ROC AUC and were used to refit each model on the entire training set.

### Anomaly-detection models and Higher-order stacking ensemble

We added three anomaly-detection models to the six main classifiers to assess the hypothesis that new T2DM incidences could be considered anomalies in the dataset; therefore, anomaly detectors may outperform classifiers. Our selected models were as follows: IF is a tree-based model that assumes anomalies lie in sparse regions of feature space and require fewer splits, thus existing in shorter branches (20). LOF is another anomaly model with a different approach; it assigns each data point a score based on the ratio of its local density compared to the average local density of its K nearest neighbors. If a point has a substantially lower score compared to its counterparts, we consider it an outlier (21). Lastly, OC-SVM creates a decision boundary that encompasses the majority of the data, regardless of its origin; observations outside of this boundary are considered outliers (22). We trained the mentioned anomaly models unsupervised on the training data and the T2DM positive class set as an anomaly.

Base learners in the stacking ensemble were the six tuned classifiers. The meta-learner was a logistic regression model with class-weight. Stacking was implemented with internal stratified 5-fold cross-validation to ensure that, for each instance, base model predictions for the meta-learner were generated using only data from folds where the instance was not included in the base model’s own training set; therefore, preventing data leakage from the meta-learner level, and predictions for each sample are never influenced by its label. The meta-learner was then retrained on the complete set of base model outputs for final testing.

### Reduced-Parameter and External Validation Models

For a head-to-head comparison between our internal cohort and external NHANES cohort, based on mapped predictors (FBS, BMI, age, sex, FH of T2DM, SESq, and CVD), 7-parameter models (micro models) were developed. Furthermore, for comparison between the internal cohort of female participants and the external PIMA Indian women cohort, we deployed 3-parameter models (age, BMI, and FBS).

### Performance evaluation

To evaluate all models on the held□out test set and external cohorts, we quantified discrimination using the Area under the receiver□operating□characteristic curve (ROC AUC) and Area under the precision□recall curve (PR AUC), since thresholds other than 0.5 are not used in clinical triage, we chose this hard□threshold and report F1□score, precision, recall, and the confusion matrix. We utilized nonparametric bootstrap confidence intervals (n = 1,000) for both probability-based metrics (ROC AUC and PR AUC) and hard-label metrics (F1, precision, and recall). Calibration was summarized using the Brier score and visually assessed with calibration curves. At each iteration, we resampled (with replacement) the test set, recomputed the metric, and took the 2.5th and 97.5th percentiles as the confidence limits. Moreover, we computed the mean difference in AUC (each model minus FINDRISC). We estimated a 95% confidence interval using a paired nonparametric bootstrap approach with 2,000 iterations to preserve the within-subject correlation in predicted probabilities in both primary and sensitivity analyses. In the sensitivity analysis, we repeated all major analytic steps identically, excluding FBS. The net benefit was calculated for each model across a range of probability thresholds (0.01– 0.99) to assess clinical utility, using both the “treat all” and “treat none” strategies in the primary internal analysis and the sensitivity analysis. 7-parameter and 3-parameter models were internally and externally (independently) evaluated on the external NHANES 2013–14 and PIMA Indian women datasets, respectively. Preprocessing and standardization for external data were performed using parameters derived solely from the internal training set to avoid information leakage. To understand the drivers of model predictions, SHAP was used. For tree-based models, we used the SHAP tree explainer. For linear models, the SHAP linear explainer was employed, and for other classifiers, the SHAP kernel explainer was applied. A random subset of 100 training instances was used as the background dataset. We computed SHAP values on a further random sample of up to 100 test subjects (for computational tractability). Finally, we ranked features by mean absolute SHAP value and displayed the top 10 in a horizontal bar plot. Further visualization included a beeswarm plot for each model and an overall comparison of feature importance across models.

### Reproducibility and Software

All analyses were performed in Python 3.12.7, using scikit-learn 1.6.1, SHAP 0.47.2, pandas 2.2.3, NumPy 1.26.4, and Seaborn, Matplotlib, and Graphviz for visualization. To advocate for open science and adhere to a policy of complete transparency, the entire code, external cohorts’ data, and the internal mock dataset with identical metrics to the real internal dataset will be made public immediately upon acceptance or publication in a GitHub repository. To incorporate this study into clinical practice, we developed a web-based clinical decision-support application based on the validated 7-parameter models. The app is publicly available at (https://t2dm-calculator.onrender.com/). The app is open-sourced, and the code is available at (https://github.com/ParsaMD/T2DM_calculator).

## Results

### Cohort description

In the internal cohort, 9171 participants were included; the mean age ± standard deviation (SD) was 46.91 years ± 8.24, and the mean follow-up time was 7.09 ± 1.25 years. Additionally, our cohort comprised 4374 (47.69%) males and 4797 (52.31%) females. Of the 9171 participants, 8904 (97.09%) entrants did not develop T2DM, and during the follow-up period, 267 new cases of T2DM were identified (Table 1a). From the NHANES cohort, 4062 participants were included, 263 of whom were diagnosed with T2DM. The mean age of non-diabetic entrants was 46.45 ± 17.25, and the mean age of diabetic attendees was 61.96 ± 13.00 (Table 1b). The PIMA cohort comprised 480 participants with a mean age of 29.72 ± 10.04 in non-diabetics and 36.07 ± 9.66 in people with diabetes (Table 1c). Utilizing our internal cohort, we included the top 16 features in the final analysis. Only the smoking variable was one-hot encoded. Since other categorical variables were either binary or had a hierarchical order (SESq), we dropped the first category (non-smokers) to avoid multicollinearity. The passive smoker category was the sole smoking status included in the final analysis (the only one in the top 16). Random forest feature importances (Figure 1 S1 file) showed that baseline FBS is by far the most important predictor, followed by BMI, waist circumference, MET, and age. In the sensitivity analysis, when the strong FBS predictor was removed, BMI, waist circumference, age, and MET were the most important predictors (Figure 10 S1 file).

### Baseline model’s performance

Initially, we performed 5-fold cross-validation on the training set. The highest mean ± SD for ROC AUC belonged to LR (0.87 ± 0.01), followed by GBM (0.86 ± 0.01), and RF (0.84 ± 0.02) (Table 3 S2 file). The 16-parameter models, including stacking, anomaly models, and FINDRISC ‘s ROC AUCs, PR AUCs, F1, and Brier scores, and their respective 95% CIs, can be found in Table 2. The full results of the internal test set, including precision and recall, are presented in Table 4 of the S2 file. In the internal cross-validation experiments, LR performed exceptionally well, achieving an ROC AUC of 0.87, which indicates that the model correctly ranks a randomly chosen instance with a probability of 87%. Followed closely by GBM (86%) and RF (84%). Twenty percent of the internal cohort dataset was held out from the beginning and used only once for the final analysis. On the test set, NN achieved the best overall discrimination (ROC AUC) (95% CI) at 0.87 (0.82-0.91), the highest PR AUC at 0.22 (0.14-032), the best precision at 0.66 (0.00-1.00), and the best uncalibrated Brier score at 0.025 (0.020-0.031); moreover, RF achieved the best balance between precision and recall (F1 score) at 0.26 (0.18-0.35) (Table 2). The stacking ensemble achieved the highest recall among the baseline models, at 0.81 (0.69-0.90). All three anomaly models had low metric values. The top model with top distinguishing power belonged to IF at 0.68 (0.62-0.75), closely followed by LOF at 0.66 (0.58-0.74). The lowest performance was observed with OC-SVM at 0.54 (0.46-0.63). PR AUC values for LOF, IF, and OC-SVM were 0.065, 0.059, and 0.041, respectively (Table 2, Figures 2 and 3). Confusion matrices, PR curves, and ROC curves for these models are presented in Figure 2 of the S1 file. Additionally, bar plots illustrating a comparison of model metrics (ROC AUC, PR AUC, F1 score, precision, and recall) are shown in Figure 3 of the S1 file. Figure 2a shows ROC AUC Forest plots with 95% CIs of all models and FINDRISC in the primary internal analysis. Almost half of the participants achieved a score between 5 and 9 (Table 1 S2 file). FINDRISC achieved a ROC AUC (95% CI) of 0.70 (0.64-0.76) and a PR AUC of 0.05 (0.03-0.09), sitting above the anomaly models and below traditional ML models and stacking. The F1 score, precision, and recall were 0.04 (0.00-0.11), 0.06 (0.00-0.17), and 0.03 (0.00-0.09), respectively. The Confusion matrix, PR curve, and ROC curve for FINDRISC are presented in Figure 4 of the S1 file. Additionally, FINDRISC metrics, including ROC AUC, PR AUC, F1 score, precision, and recall, are compared with other models in Table 4 of the S2 file. Table 3 shows paired bootstrap mean differences in ROC AUC (with 95% confidence intervals) compared to FINDRISC. All classifiers and stacking outperformed FINDRISC in the main internal analysis. All decision curve analysis (DCA) curves for our models in internal main and sensitivity analysis were flat at zero net benefit, indicating that at no threshold was there a positive clinical benefit to using the model compared with either treating all or none (see Figure 5 for main analysis and Figure 13 for sensitivity analysis in the S1 file).

**Table 3.** Paired bootstrap mean differences in ROC AUC (with 95% confidence intervals) for each machine learning model compared to FINDRISC in primary and sensitivity analyses.

| **Model (vs FINDRISC)** | **Mean AUC diff (Primary Analysis)** | **95% CI** | **Mean AUC diff (Sensitivity Analysis)** | **95% CI** |
| --- | --- | --- | --- | --- |
| **Neural Net** | 0.17 | 0.10 - 0.23 | -0.05 | -0.14 - 0.02 |
| **Stacking Ensemble** | 0.16 | 0.10 - 0.23 | 0.01 | -0.04 - 0.08 |
| **Random Forest** | 0.15 | 0.09 - 0.22 | 0.01 | -0.05 - 0.08 |
| **Decision Tree** | 0.15 | 0.08 - 0.23 | -0.01 | -0.10 - 0.06 |
| **SVM** | 0.14 | 0.08 - 0.21 | 0.03 | -0.01 - 0.08 |
| **Logistic Regression** | 0.14 | 0.08 - 0.20 | 0.00 | -0.04 - 0.06 |
| **Gradient Boosting** | 0.14 | 0.07 - 0.21 | 0.02 | -0.02 - 0.09 |
| **Isolation Forest** | -0.01 | -0.07 - 0.04 | -0.05 | -0.12 - -0.00 |
| **Local Outlier Factor** | -0.03 | -0.11 - 0.04 | -0.08 | -0.15 - -0.01 |
| **One Class SVM** | -0.15 | -0.24 - -0.06 | -0.17 | -0.26 - -0.08 |

**Figure 2.**
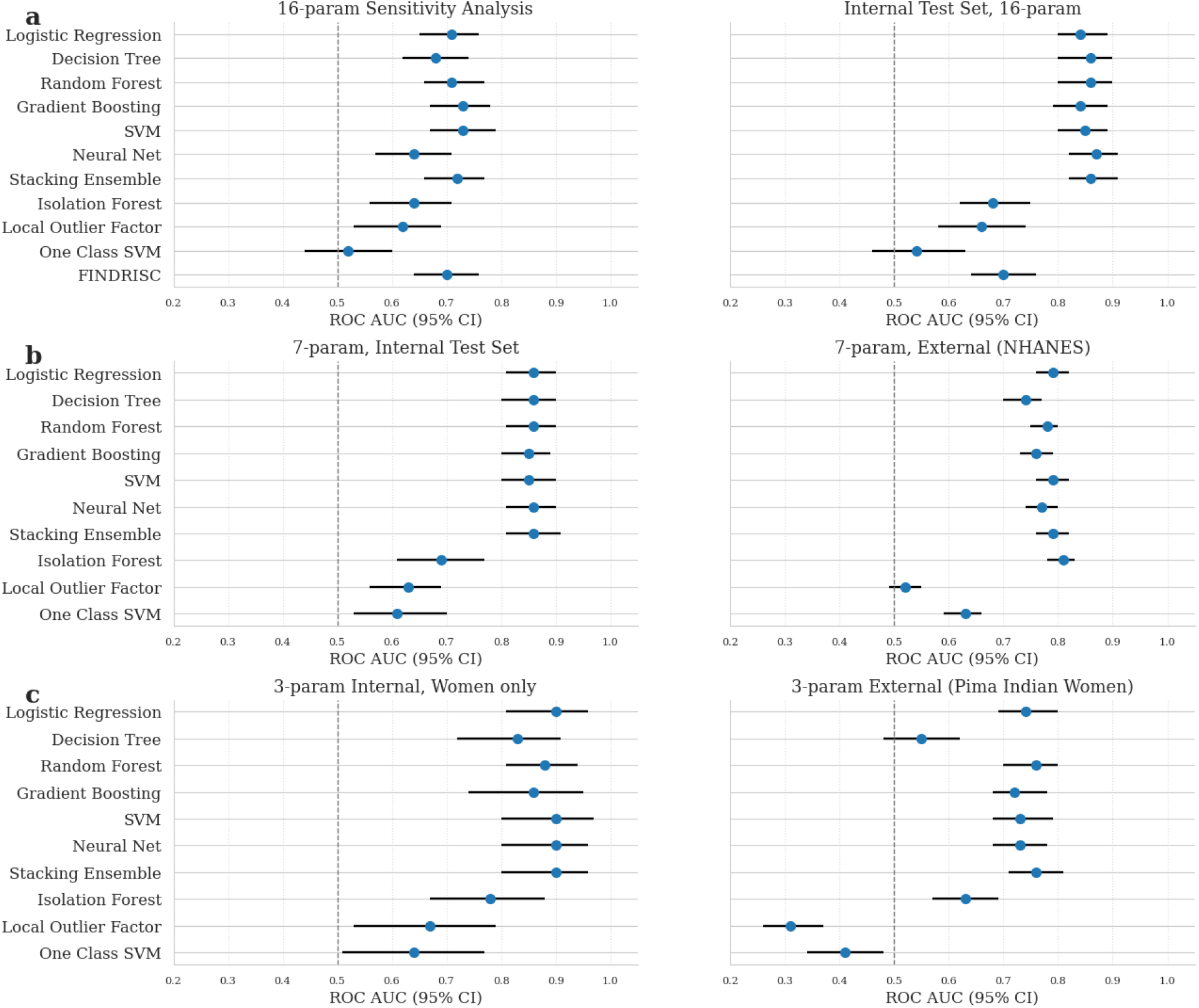
ROC AUC Forest Plot with 95% Confidence Intervals. (a) 16-variable models: sensitivity analysis (without fasting blood sugar, left) and complete main internal test set (right). (b) 7-variable models: internal test set (left) and external NHANES validation (right). (c) 3-variable models: internal test set (women only, left) and external Indian Pima cohort (right).

**Fig 3.**
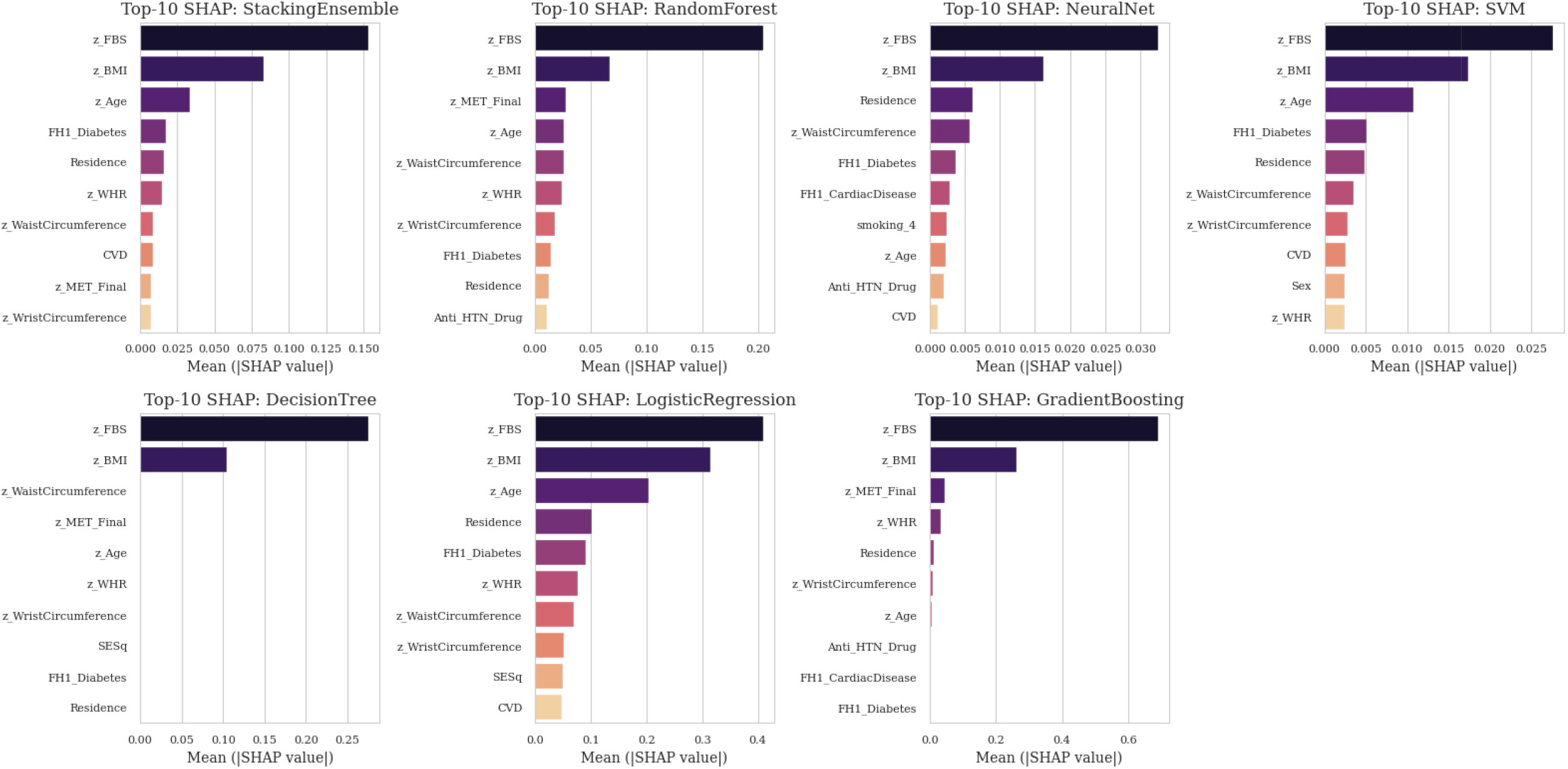
Global explainability via SHAP (main test set, 16-parameter models): top-10 SHAP barplot for stacking and six classifiers.

### Sensitivity analysis

As it is evident that FBS is the strongest predictor, we performed a sensitivity analysis to see the models’ performance without FBS. We repeated the entire modeling pipeline exactly as before but dropped FBS from the feature set. Table 5 of the S2 file summarizes the performance of all models (ROC AUC, PR AUC, F1, precision, and recall), each with 95% bootstrap CIs on the same test split. All models showed lower performance across metrics in the sensitivity analysis compared to the primary analysis. In the main analysis, all classifiers achieved ROC AUCs above 0.84; however, in the sensitivity analysis, the best-performing model was SVM, with a ROC AUC (95% CI) of 0.736 (0.679–0.792), closely followed by GBM (0.731, 0.676–0.783) and stacking (0.72, 0.66–0.77). FINDRISC, as in the primary analysis, performed below all traditional ML models (ROC AUC 0.70), yet in the sensitivity analysis, it outperformed NN and DT for discrimination (Table 2a and Figure 2a). Furthermore, anomaly detection models also had slightly lower ROC AUCs (IF: 0.64 (0.56-0.71), LOF: 0.62 (0.53-0.69), and OC-SVM: 0.52 (0.44-0.60)) compared to the main analysis. PR AUCs, F1, precision, and recall values were also lower under the sensitivity analysis. Table 3 presents paired bootstrap mean differences in ROC AUC (with 95% confidence intervals) compared to FINDRISC under the sensitivity analysis condition. No model statistically significantly outperformed FINDRISC in the sensitivity analysis (all 95% CIs included zero). Confusion matrices, PR curves, and ROC curves for all models under sensitivity analysis conditions are presented in Figure 11 of the S1 file. Additionally, bar plots illustrating a comparison of model metrics are shown in Figure 12 of the S1 file.

### Calibration analysis

Model calibration was evaluated using the Brier score and calibration plots. As shown in Table 2d, all models achieved generally low Brier scores, indicating reasonable average accuracy of their predicted probabilities. Among them, SVM, GBM, and NN had the lowest Brier scores. Both simple models (such as SVM) and more complex models (such as NN) showed good calibration. Visual inspection of the calibration curves confirmed that these models were the best calibrated. Calibration plots for all classifiers in the primary and sensitivity analyses are shown in Figure 9 and Figure 17 of the S1 file. All Brier scores and calibration curves are based on uncalibrated predicted probabilities.

### Reduced Models and External Validation

We were able to map seven features of the NHANES 2013-2014 with our internal cohort (Table 2 S2 file); furthermore, for an honest and fair comparison, we retrained all models and created 7-parameter models (micro models) and validated them externally and temporally on a different ethnicity using the NHANES dataset. On the internal test set RF with ROC AUC (95% CI): 0.86 (0.81-0.90), stacking: 0.86 (0.81-0.91), and LR: 0.86 (0.81-0.90) were the best performing models, on the validation NHANES dataset LR with ROC AUC (95% CI): 0.79 (0.76-0.82), stacking: 0.79 (0.76-0.82), and SVM: 0.79 (0.76-0.82) were the best ML classifiers. Surprisingly, the IF anomaly performed exceptionally well on the NHANES dataset, with a ROC AUC (95% CI) of 0.81 (0.78-0.83). For a visual comparison of models’ performance using ROC AUC, Forest plots with 95% CIs are shown in Figure 2b. Additionally, Table 2 displays the differences in ROC AUC, PR AUC, F1 score, and Brier score between internal 7-parameter and external 7-parameter models. For a complete comparison using other metrics, such as precision and recall, please refer to Tables 6 and 7 in the S2 file.

To further assess the generalizability of our models and to perform sex-specific and subgroup analyses, we utilized the publicly available PIMA Indian women cohort data. The PIMA dataset contains only a limited set of features; therefore, to fairly evaluate our models under conditions of feature scarcity and to examine model performance with a highly parsimonious approach, we created 3-parameter models (nano models), and tested them on the internal test set (only women) and the PIMA dataset. In the internal test set, LR with ROC AUC (95% CI): 0.90 (0.81-0.96), stacking 0.90 (0.80-0.96), and SVM: 0.90 (0.80-0.97), indicating excellent discrimination. On the PIMA dataset, stacking 0.76 (0.71-0.81), RF 0.76 (0.70-0.80), and LR: 0.74 (0.69-0.80) yielded the highest ROC AUCs. Notably, for the first time in our evaluation, two anomaly detection models demonstrated ROC AUC values below 0.50 on the PIMA cohort: LOF scored 0.31 (0.26–0.37) and OC-SVM 0.41 (0.34–0.48). Figure 2c shows ROC AUC Forest plots with 95% CIs of nano models; additionally, Table 2 exhibits ROC AUCs, PR AUCs, F1, and Brier scores of the mentioned models. The complete results of the nano models on the internal test set and the PIMA dataset are presented in Tables 8 and 9 of the S2 file, respectively.

An overall view of the Best-performing models, as indicated by PR AUC, F1, precision, and recall across cohorts, is presented in Table 10 of the S2 file. Confusion matrices, PR curves, and ROC curves for the internal 7-parameter, external 7-parameter, internal 3-parameter, and external 3-parameter models are shown in Figure 18, Figure 19, Figure 20, and Figure 21 of the S1 file, respectively.

### Interpretability

To understand the contribution of each feature to model predictions, we performed the SHAP test across all ML models. Figure 3 shows the mean absolute SHAP values for the top 10 features in each model. We used a random sample from the entire cohort, which allowed us to compare the influences of features directly and provided insight into the decision-making processes of each model. Baseline FBS was the most influential predictor across all models, per clinical relevance in T2DM risk. Other features frequently ranked among strong predictors were BMI and age. Tree-based models (RF, DT, and GBM) emphasized FBS and BMI for predicting the outcome, whereas LR, SVM, NN, and stacking assigned importance to broader predictors, such as waist circumference, age, and residence. Figure 6 of the S1 file shows the top-10 global SHAP feature importances of all 16-parameter models in the primary analysis. Figure 7 of the S1 file displays a cross-model SHAP feature importance plot for all models included in the main analysis. Next to strong features like FBS, BMI, and age, residence type, FH of T2DM, and WHR are the most important ones. Figure 8 (a-j) of the S1 file displays SHAP beeswarm plots for the 16-parameter models, revealing that FBS, BMI, and age have the most significant and most consistent contributions to model predictions, with additional contributions from residence type, family history of type 2 diabetes, and waist-hip ratio. We repeated the same approach in the sensitivity analysis. Figure 14 of the S1 file shows the top-10 global SHAP feature importances of all sensitivity analysis models. BMI and age remain the most influential predictors, followed by FH of T2DM, WHR, and residence type. Figure 15 of the S1 file displays a cross-model SHAP feature importance plot for all models in the sensitivity analysis, presented in the same order as the features, which is validated here again. Figure 16 (a-j) of the S1 file shows SHAP beeswarm plots for the 16-parameter models under sensitivity analysis conditions.

## Discussion

### Principal Findings

In this study, we benchmarked six ML classifiers, a stacking ensemble, and three anomaly detectors against FINDRISC in predicting incident T2DM in a large prospective cohort. Our rigorous pipeline (data harmonization, feature selection, cross-validation, bootstrapping, calibration, DCA, SHAP, external validation) enabled us to compare comprehensively ML vs. clinical scores. In the main analysis, ML classifiers outperformed FINDRISC (AUC 0.70, 95% CI 0.64–0.76). NN achieved an ROC AUC of 0.87 (95% CI 0.82– 0.91), and RF/stacking achieved 0.86, with a stacking recall of 0.81, which would reduce missed cases if deployed in practice. We also wanted to check if baseline FBS is not available (simulating a non-laboratory, questionnaire-only scenario), whether ML models could outperform FINDRISC again. In the setting mentioned above, the ROC AUC of all ML models decreased by ≥0.14. FINDRISC then outperformed NN, with SVM performing best among ML classifiers (ROC AUC 0.73, 95% CI 0.67–0.79). This highlights the continued value of FINDRISC in non-lab settings and ML reliance on lab features; ML in these settings matches but does not surpass FINDRISC.

We provide, for the first time, paired head-to-head comparisons with external validation, bootstrapped CIs, DCA, and robust calibration. SHAP analyses indicated that baseline FBS, BMI, waist circumference, and age were among the most influential predictors across models. The interpretability analysis also sheds light on the black box concerns hindering ML clinical acceptance. A recent study by Lugner et al. yielded similar results in identifying the best predictors of new T2DM incidence; BMI, waist circumference, glucose levels, and age were among the top predictors. In contrast, they did not provide results excluding laboratory-associated features (6). In a meta-analysis, De Silva et al. included 23 studies that applied ML tools to predict T2DM incidence; however, none of these studies tested anomaly models or externally validated their models against FINDRISC (23). Another systematic review compared ML models with each other; they found that tree-based models (RF, DT, and GBM) perform extraordinarily well in terms of ROC AUC. Additionally, they interpreted that lifestyle and socioeconomic features, coupled with laboratory-based features, could substantially predict T2DM incidence, consistent with our conclusion (24).

### External Validation

All ML classifiers demonstrated a decline in ROC AUC upon both external validations; several factors likely contributed to this drop. Covariate shift and population heterogeneity may be the first reasons that predictor-outcome relationships, baseline risks, and covariate distributions differ between internal and external cohorts. Furthermore, not all variables could be mapped identically (e.g., SES, CVD definitions, and the settings in which blood samples were acquired for FBS varied across cohorts). Micro and nano models necessarily omitted some top-ranked features as identified by internal SHAP analysis. Moreover, we intentionally avoided model refitting or recalibration on external data to provide an honest test of transportability. This strict validation approach generally yields lower but more realistic estimates of external performance.

By having an imbalanced dataset, we hypothesized that the positive class could be considered an anomaly in the dataset; therefore, we utilized anomaly detection models. Unfortunately, unsupervised anomaly detection models proved to be useless in the main and sensitivity analysis, with IF emerging as the best in terms of ROC AUC 0.68 (95% CI 0.62–0.75) in the main analysis and 0.64 (0.56-0.71) in the sensitivity analysis, underperforming FINDRISC in both scenarios. Nonetheless, in NHANES external validation, anomaly detection models, especially IF (ROC AUC: 0.81, 95% CI: 0.78-0.83), performed better than in the internal cohort and outperformed all the classifiers. This improvement likely reflects their insensitivity to label distribution shifts; with fewer predictors and consistent data structure, identifying T2DM as an outlier became more effective. However, despite this gain, anomaly models still did not surpass the top supervised methods elsewhere and lack interpretability or calibration for clinical use.

### Clinical Utility & DCA

Despite the strong discriminative performance of several ML models, DCA across all settings demonstrated consistently flat net benefit curves near zero (see Figure 5 and Figure 13 in S1), regardless of threshold or modeling approach. This finding persisted in both the primary and sensitivity analyses. This flatness can be attributed primarily to the extreme class imbalance in the internal cohort (T2DM incidence: 2.9%), which leads to an excessive number of true negatives and an inherently low absolute number of true positives and false positives. In such settings, even well-calibrated models that statistically distinguish between cases and non-cases may yield a limited clinical net benefit. Uncalibrated Brier scores remained acceptable in ML classifiers across all cohorts (0.02-0.27), indicating that probability outputs were well-aligned with observed outcomes.

In practical terms, the addition of ROC AUC from 0.70 (FINDRISC) to almost 0.87 (NN, stacking) and the 0.81 recall achieved by our stacking, if adopted as a clinical prediction tool, translates to fewer missed T2DM cases. However, F1 scores were not substantially high due to an extremely imbalanced dataset; additionally, there was a harsh trade-off between precision and recall, as models either acted too conservatively to increase precision or reported too many false positives. The paired bootstrap analysis (Table 3) indicates that ML classifiers can significantly improve upon the discriminative performance of the FINDRISC score; however, this finding was not robust under sensitivity analysis conditions. The lack of statistically significant improvement in the sensitivity analysis shows the importance of features to ML models, and in our case, the importance of baseline FBS in predicting T2DM incidence using ML models.

### Limitations

In this project, we faced several limitations. First, we didn’t include the dietary habits part in our FINDRISC score calculations and assigned a zero score in that domain. Secondly, we wanted to conduct DeLong test to compare ROC AUC of the models, but due to extreme imbalance and small number of predicted positive cases, we were unable to draw meaningful results; nevertheless, we mitigated this limitation by using paired bootstrap confidence intervals, which provide a robust estimation of uncertainty even in datasets with limited event rates. Third, the NHANES dataset was a cross-sectional dataset, and instead of T2DM incidence, we used T2DM prevalence as the outcome of interest.

### Future Work

Strengths include a large and diverse cohort, a rigorous pipeline, internal and external validation, and explainability. Our public code offers a reproducible workflow. Furthermore, clinicians can use the web calculator for experimental use in clinics. Future research should pursue multi-site validation, incorporate additional data sources (e.g., genomics), and enhance translation through user-friendly tools, such as our online calculator.

## Conclusion

In conclusion, when provided with appropriate laboratory data, ML models can deliver substantial gains in T2DM risk stratification over established clinical scores. In resource-limited or survey-only settings, FINDRISC performs well. Transparent, calibrated, and externally validated analytics, as demonstrated here, are essential for the responsible clinical adoption of ML-based diabetes prediction.

## Data Availability

The code used for analysis will be made available upon publication or acceptance.

## Ethical approval

Ethical approval for the original Ravansar Non-Communicable Disease (RaNCD) cohort study was granted by the Ethics Committee of Kermanshah University of Medical Sciences (approval code: KUMS.RE.1401.134). All procedures involving study participants were conducted by relevant institutional and national guidelines and regulations. Written and oral informed consent was obtained from all participants before enrollment in the cohort. This secondary analysis utilized anonymized data from the RaNCD study, with all identifying information removed prior to analysis. The NHANES and PIMA datasets were publicly available.

## Funding

None.

## Competing interests

The authors declare that they have no conflict of interest.

## Code availability

The code used for analysis will be made available upon publication or acceptance.

## Authors’ contributions

Parsa Amirian:

Study design, data collection, data analysis, data interpretation, and writing the original manuscript.

Mahsa Zarpoosh:

Data collection, data interpretation, writing the original manuscript, revision, and figures

## Notes

### Competing Interest Statement

The authors have declared no competing interest.

### Funding Statement

This study did not receive any funding

